# Incorporating Nonlinear Kinetics to Improve the Predictive Performance of Population Pharmacokinetic Models for Ciclosporin in Adult Renal Transplant Recipients: A Comparison of Modelling Strategies

**DOI:** 10.1101/2020.01.25.20018820

**Authors:** Jun-Jun Mao, Zheng Jiao, Xiao-Yan Qiu, Ming Zhang, Ming-Kang Zhong

## Abstract

**Aim:** Ciclosporin (CsA) has been shown to follow nonlinear pharmacokinetics in renal transplant recipients who received Neoral-based triple immunosuppressive therapy. Some of these nonlinear properties have not been fully considered in population pharmacokinetic (popPK) analysis. Therefore, the aim of this study was to determine the potential influence of nonlinearity and the functional forms of covariates on model predictability.

**Methods:** A total of 2969 CsA whole-blood measurements, including 1328 pre-dose and 1641 2-h post-dose concentrations, were collected from 173 patients who underwent their first renal transplantation. Four popPK models based on different modelling strategies were developed to investigate the discrepancy between empirical and theory-based, linear and nonlinear compartmental kinetic models and empirical formulae on model predictability. Prediction-based and simulation-based diagnostics (prediction-corrected visual predictive checks) were performed to determine the stability and predictive performance of these four models.

**Results:** Model predictability improved when nonlinearity was considered. The theory-based nonlinear model which incorporated nonlinear property based on known theoretical relationships performed better than the other two compartmental models. The nonlinear Michaelis-Menten model showed a remarkable improvement in predictive performance over that of the other three compartmental models. The saturated binding of CsA to erythrocytes, and auto-inhibition that arose from the inhibitory effects of CsA on *CYP3A4*/P-gp and CsA-prednisolone drug interaction may have contributed to the nonlinearity.

**Conclusions:** Incorporating nonlinear properties are likely to be a promising approach for improving CsA model predictability. However, CsA nonlinear kinetics resources need further investigation. Until then, Michaelis-Menten empirical model can be used for CsA dose adjustments.

**What is already known about this subject:**

- CsA in renal transplant recipients receiving Neoral-based triple immunosuppressive therapy followed nonlinear pharmacokinetics.
- Nonlinearity is rarely incorporated into CsA population pharmacokinetic (popPK) modelling processes.

**What this study adds:**

- Four popPK models based on different modelling strategies were developed to investigate the discrepancy between empirical and theory-based compartmental kinetic models and empirical formulae, as well as the effect of nonlinearity on CsA model predictability.
- Based on the four models, incorporating nonlinear properties is likely to be a promising approach for improving CsA model predictability.
- Saturated distribution into red blood cells, and auto-inhibition that arose from the inhibitory effects of CsA on *CYP3A4*/P-gp and CsA-prednisolone drug interaction may be the main sources of CsA PK nonlinearity.

**Principal Investigator statement:** The authors confirm that the Principal Investigator for this paper is Zheng Jiao and that he had direct clinical responsibility for patients.

## 1 Introduction

Ciclosporin (CsA), an immunosuppressant with a narrow therapeutic range and considerable pharmacokinetic (PK) variability, is widely used to prevent allograft rejection after renal transplantation [1, 2]. CsA has low oral bioavailability (approximately 25%, range 10% - 89%) [3] due to its erratic gastrointestinal absorption and the combined activity of cytochrome P450 (CYP) 3A and P-glycoprotein (P-gp) [1]. It is extensively bound to erythrocytes [3], metabolised by *CYP3A4* and *CYP3A5* [4], and then mainly eliminated in bile [5, 6].

The absorption and disposition of conventionally formulated CsA are considered dose-dependent [7]. To reduce oral absorption variability and improve CsA dose linearity, a new oral microemulsion of CsA (Neoral^®^, Novartis) was formulated [8]. Mueller *et al*. [9] compared the PK of Neoral with that of the conventional CsA formulation and concluded that Neoral had superior dose linearity with exposure after oral administration. However, it should be noted that this study was conducted in healthy volunteers administered a single oral dose.

In contrast to single-dose PK studies, the in-vivo behaviour of drugs in clinical practice is co-influenced by multiple intrinsic/extrinsic factors including demographic factors, genetic polymorphisms of drug metabolising enzymes and transporters, disease progression, concomitant medications, and most importantly their combined effects [10]. As determined by population pharmacokinetic (popPK) analyses, CsA is revealed to have a nonlinear PK in renal transplant recipients [11, 12].

The observed nonlinearity phenomenon may be partially attributed to metabolic enzyme/transporter protein interactions. In addition to CYP3A and P-gp substrates, CsA potently inhibits transporter proteins P-gp and OATP1B1, as well as CYP3A4 [13-16]. Additionally, CsA is typically used in combination with anti-proliferative drugs and corticosteroids in clinical settings [17]. When co-administered with steroids, it is able to activate pregnane X receptor (PXR), and then up-regulates *CYP3A* and *MDR1* gene expression [18-20]. CsA-prednisolone drug interaction may cause CsA PK nonlinearity. Furthermore, the saturated binding of CsA to erythrocytes may also contribute to nonlinearity [3].

Obtaining an accurate description of the complex PK processes of CsA is challenging but useful in supporting dosage adaptation. Numerous studies have described the popPK characteristics of CsA in adult renal transplant recipients [11, 21]. The available models have identified covariates mainly through empirical investigation. No consistent covariates or their functional forms in models were identified in those studies. Some of the nonlinear properties described above are not fully considered in modelling processes. Including covariate effects based on theoretical mechanisms rather than empirically identifying relationships should lead to more consistent results [22].

To determine the potential influence of nonlinearity and the functional forms of covariates on model predictability, four popPK models based on different modelling strategies were developed. These models investigated the discrepancy between empirical and theory-based compartmental kinetic models and empirical formulae, as well as the effect of nonlinearity on model predictability. Their predictive performance was evaluated by prediction- and simulation-based diagnostics, as previously reported [11, 23].

## 2 Materials and Methods

### 2.1 Patients

Data from 173 adults (112 male and 61 female), who underwent their first renal transplantation at Huashan Hospital were retrospectively collected. Patients were randomly divided into two groups: 121 for model development and 52 for model evaluation. Whole-blood samples of CsA pre-dose concentration (C_0_) and 2-h post-dose concentration (C_2_) were monitored. Demographic and pathophysiological data were obtained during routine clinical visits from July 2003 to December 2016. Patient follow-up was conducted up to three years after surgery. Those undergoing dialysis treatments were excluded from this study. The observations from the first two postoperative days were excluded to ensure that the measured CsA concentrations were relatively stable.

The study protocols were approved by the Ethics Committee of Huashan Hospital and written informed consent was obtained from all subjects.

### 2.2 Immunosuppressive Therapy

All patients received combined immunosuppressive therapy comprising a CsA microemulsion (Neoral^®^; Novartis Pharma Schweiz AG, Emberbach, Germany), mycophenolate mofetil (MMF, CellCept^®^; Roche Pharma Ltd, Shanghai, China) and corticosteroids. Initial CsA dosage was 5 mg kg^-1^ day^-1^, administered as two doses under fasting conditions immediately after surgery. CsA concentration were regularly monitored to adjust the dosage and achieve target concentrations based on local guidelines (Text S1) [24]. MMF (0.5 - 3 g day^-1^) was administered based on body weight (WT) and postoperative days (POD). This schedule was followed by oral prednisolone (80 mg day^-1^) and the dosage was gradually decreased by 10 mg day^-1^ until reaching 20 mg day^-1^ on day 10. The dosage was further tapered to 15, 10, and 5 mg day^-1^ by months 1, 3, and 6, respectively.

### 2.3 Bioassay

Whole blood samples were treated with the anti-coagulant ethylene diamine tetraacetic acid and analysed with a well-validated fluorescence polarisation immunoassay (FPIA) using an AxSYM^®^ Abbott diagnostic system (Abbott Diagnostics, Chicago, IL, USA) between July 2003 and April 2011, and a chemiluminescent microparticle immunoassay (CMIA) using an Architect I2000 system between May 2011 and Deccember 2016. For AxSYM, the limit of detection (LOD) was 21.8 ng mL^-1^, and the calibration range was 40 - 800 ng mL^-1^; for CMIA, the LOD was 25 ng mL^-1^, and the calibration range was 30-1500 ng mL^-1^.

The following formulae (Eq. 1) [25] was used to convert CMIA measured C_0_ before modelling.

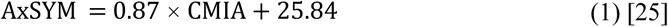

where AxSYM represents the FPIA performed using an AxSYM analyser and CMIA represents the chemiluminescent microparticle immunoassay.

### 2.4 Population Pharmacokinetic Model Development

Population PK analysis was performed using the NONMEM^®^ software package (version 7.4; ICON Development Solutions, Ellicott City, MD, USA), with Pirana^®^ 2.9 as an interface for Perl Speaks NONMEM (PsN; version 4.9.0) and R software (version 3.5.0, http://www.r-project.org/) [26]. The first-order conditional estimation method including η-ε interaction (FOCE-I) was used throughout the method-building procedure [27].

#### 2.4.1 Base Model

To determine the potential influence of nonlinearity and the functional forms of covariates on model predictability, four popPK models (linear compartmental model without incorporating daily dose, linear compartmental model incorporating daily dose, theory-based nonlinear compartmental model, and nonlinear Michaelis-Menten (MM) empirical model) were developed based on two structural models.

Since only the therapeutic drug monitor (TDM) C_0_ and C_2_ concentrations were available for analysis, one-compartmental model with first-order absorption was used to describe CsA PKs in compartmental (CMT) models. The model was parameterised in terms of apparent total clearance (CL/*F*), apparent volume of distribution (V/*F*), and the absorption rate constant (K_a_). K_a_ was fixed at 1.25 h^-1^ based on literature values [11, 28].

A nonlinear MM empirical formula (Eq. 2) that quantified the overall relationship between daily dose and CsA concentrations was used as a structural model to describe CsA PKs in the MM model [11].

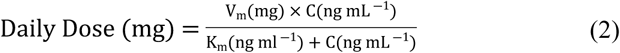

where V_m_ denotes the maximum dose rate (daily dose), K_m_ denotes the steady-state concentrations at half-maximal dose rate and C represents the CsA C_0_ or C_2_ concentrations.

Between subject variability (BSV) was assumed to be log-normally distributed, and estimated for all parameters, except K_a_. Residual unexplained variability (RUV) was described by testing proportional, combined proportional, and additive structures.

#### 2.4.2 Covariate Model

The demographic and pathophysiological data, as well as concomitant medications (Table 1), were evaluated as covariates. Information extracted from the medical records including body size, haematocrit, postoperative days (PODs), CsA daily dose, and prednisolone dose were evaluated as the possible covariates of CsA PK. These covariates were selected according to the previous report and clinical relevance [11].

**Table 1.**
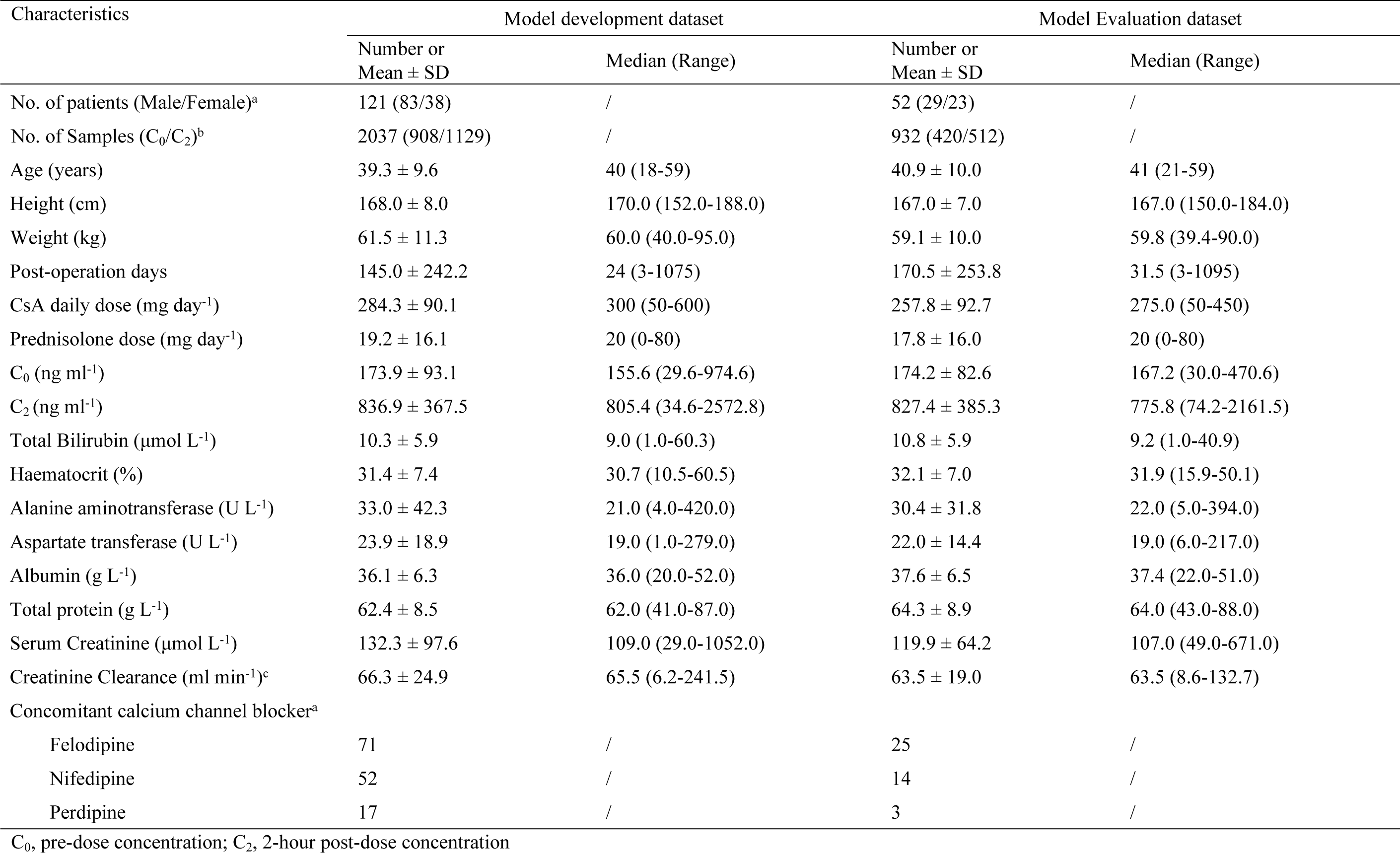
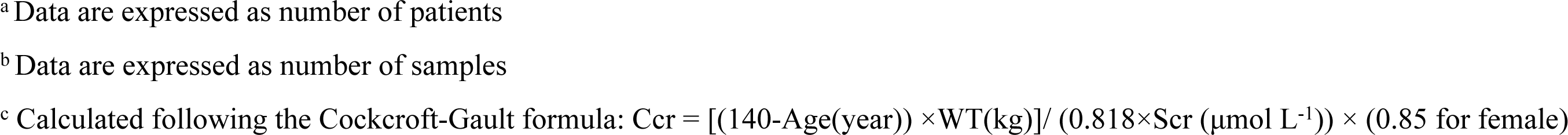
Patients demographics used to develop and evaluate population model.

| Characteristics | Model development dataset |  | Model Evaluation dataset |  |
| --- | --- | --- | --- | --- |
| | Number or Mean ± SD | Median (Range) | Number or Mean ± SD | Median (Range) |
| No. of patients (Male/Female)<sup>a</sup> | 121 (83/38) | / | 52 (29/23) | / |
| No. of Samples (C<sub>0</sub>/C<sub>2</sub>)<sup>b</sup> | 2037 (908/1129) | / | 932 (420/512) | / |
| Age (years) | 39.3 ± 9.6 | 40 (18-59) | 40.9 ± 10.0 | 41 (21-59) |
| Height (cm) | 168.0 ± 8.0 | 170.0 (152.0-188.0) | 167.0 ± 7.0 | 167.0 (150.0-184.0) |
| Weight (kg) | 61.5 ± 11.3 | 60.0 (40.0-95.0) | 59.1 ± 10.0 | 59.8 (39.4-90.0) |
| Post-operation days | 145.0 ± 242.2 | 24 (3-1075) | 170.5 ± 253.8 | 31.5 (3-1095) |
| CsA daily dose (mg day<sup>-1</sup>) | 284.3 ± 90.1 | 300 (50-600) | 257.8 ± 92.7 | 275.0 (50-450) |
| Prednisolone dose (mg day<sup>-1</sup>) | 19.2 ± 16.1 | 20 (0-80) | 17.8 ± 16.0 | 20 (0-80) |
| C<sub>0</sub> (ng ml<sup>-1</sup>) | 173.9 ± 93.1 | 155.6 (29.6-974.6) | 174.2 ± 82.6 | 167.2 (30.0-470.6) |
| C<sub>2</sub> (ng ml<sup>-1</sup>) | 836.9 ± 367.5 | 805.4 (34.6-2572.8) | 827.4 ± 385.3 | 775.8 (74.2-2161.5) |
| Total Bilirubin (μmol L<sup>-1</sup>) | 10.3 ± 5.9 | 9.0 (1.0-60.3) | 10.8 ± 5.9 | 9.2 (1.0-40.9) |
| Haematocrit (%) | 31.4 ± 7.4 | 30.7 (10.5-60.5) | 32.1 ± 7.0 | 31.9 (15.9-50.1) |
| Alanine aminotransferase (U L<sup>-1</sup>) | 33.0 ± 42.3 | 21.0 (4.0-420.0) | 30.4 ± 31.8 | 22.0 (5.0-394.0) |
| Aspartate transferase (U L<sup>-1</sup>) | 23.9 ± 18.9 | 19.0 (1.0-279.0) | 22.0 ± 14.4 | 19.0 (6.0-217.0) |
| Albumin (g L<sup>-1</sup>) | 36.1 ± 6.3 | 36.0 (20.0-52.0) | 37.6 ± 6.5 | 37.4 (22.0-51.0) |
| Total protein (g L<sup>-1</sup>) | 62.4 ± 8.5 | 62.0 (41.0-87.0) | 64.3 ± 8.9 | 64.0 (43.0-88.0) |
| Serum Creatinine (μmol L<sup>-1</sup>) | 132.3 ± 97.6 | 109.0 (29.0-1052.0) | 119.9 ± 64.2 | 107.0 (49.0-671.0) |
| Creatinine Clearance (ml min<sup>-1</sup>)<sup>c</sup> | 66.3 ± 24.9 | 65.5 (6.2-241.5) | 63.5 ± 19.0 | 63.5 (8.6-132.7) |
| Concomitant calcium channel blocker<sup>a</sup> |  |  |  |  |
| Felodipine | 71 | / | 25 | / |
| Nifedipine | 52 | / | 14 | / |
| Perdipine | 17 | / | 3 | / |

C<sub>0</sub>, pre-dose concentration; C<sub>2</sub>, 2-hour post-dose concentration
<sup>a</sup> Data are expressed as number of patients
<sup>b</sup> Data are expressed as number of samples
<sup>c</sup> Calculated following the Cockcroft-Gault formula: $Ccr = [(140 - \text{Age}(\text{year})) \times \text{WT}(\text{kg})] / (0.818 \times \text{Scr} (\mu\text{mol L}^{-1})) \times (0.85 \text{ for female})$

As reviewed previously [11], weight was the most frequently identified covariate in the final models. Therefore, PK disposition parameters were firstly related to body size based on allometric scaling theory [29, 30]. Body size was based on fat free mass predicted from total body weight, height and sex (Text S2) [31].

The potential influence of nonlinearity and the functional forms of covariates on model predictability was tested using four modelling strategies.

##### Strategy I: Linear compartmental model without incorporating daily dose

As the influence of body size was considered, covariates other than body size were investigated empirically through linear, piecewise linear, power, exponential or sigmoid functions in *Strategy I*. The concomitant medications were evaluated as binary covariates by estimating the parameter fractional change in one group compared to the other.

##### Strategy II: Linear compartmental model incorporating daily dose

Based on the result of *Strategy I*, CsA daily dose was empirically investigated on CL/*F* in this modelling strategy.

##### Strategy III: Theory-based nonlinear compartmental model

The theory-based nonlinear compartmental model was developed based on well-accepted theoretical relationships. CsA is extensively distributed in peripheral tissues and binds to erythrocytes and plasma proteins [3, 32, 33]. PK disposition parameters were estimated from the model-predicted plasma concentrations (C_p_) rather than measured whole-blood concentrations (C_wb_) under the assumption that the binding of CsA to plasma components is linear and the binding to erythrocytes is nonlinear [32, 34]. Similar to descriptions by Størset *et al*.[35] and Van Erp *et al*.[36], the following equation was used to estimate CsA C_p_ (Text S3) [37]:

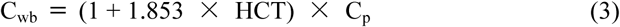

where C_wb_ is the whole-blood concentration, C_p_ is the plasma concentration, and HCT is the haematocrit level.

Therefore, all parameter estimates are expressed as plasma PK parameters in *Strategy III* and the model is capable of predicting both whole-blood and plasma concentrations.

Corticosteroids are known inducers of CYP3A enzymes and P-glycoprotein in the small intestine and/or liver [19, 20] and, theoretically, lead to CsA CL/*F* alteration. Considering the hypothesis that tapering prednisolone dosage (Pred) is the source of CsA nonlinearity, a sigmoid E_max_ model was implemented to describe the effect of prednisolone induction on CsA PKs.

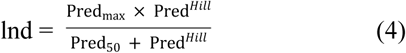

The term lnd in Eq. 4 represents the induction function relating to Pred. Pred_max_ is the maximum change in the PK parameter of interest (i.e. *F* or CL), Pred_50_ is the prednisolone dose causing half maximum induction and Hill is the sigmoid shape coefficient.

##### Strategy IV: Nonlinear Michaelis-Menten empirical model

After body size was included based on allometric scaling theory, the influence of other covariates on the MM constant (K_m_) were investigated empirically. As the CsA steady-state PK changed with time after transplantation, time factors (A/θ, Eq. 5) were introduced to test whether the MM model with time-variant K_m_ was superior [12].

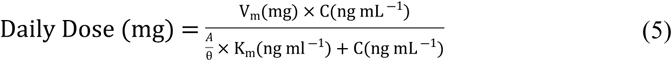

Where A was POD if 1 ≤ POD ≤ θ, or A was θ if POD > θ, V_m_ denotes the maximum dose rate (daily dose), K_m_ denotes the steady-state concentrations at half-maximal dose rate and C represents the CsA C_0_ or C_2_ concentrations.

#### 2.4.3 Model Selection Criteria

The log-likelihood ratio test was the main model selection tool. Change in objective function value (ΔOFV) was calculated to assess the statistical significance of each covariate and to differentiate between theoretically equivalent models [27]. Statistical significance levels of *p* < 0.01 and *p* < 0.001 (Chi-square distribution) were implemented in forward inclusion and backward elimination procedures, respectively. Moreover, a clear pharmacological or biological basis was also considered when covariates were included.

### 2.5 Model Evaluation

Fifty-two patients from the evaluation group were used to examine the predictability of the final four models. Estimated population predictions (PREDs) were compared to the corresponding observations (OBS) using the MAXEVAL=0 option (Eq. 6). The accuracy and imprecision of predictive performance were investigated using median prediction error (MDPE) and median absolute prediction error (MAPE), respectively [38].

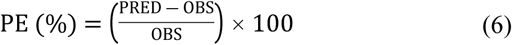

The percentage of |PE| within 20% (F20) and 30% (F30) was also calculated. The model with lower MDPE and MAPE values and fewer prediction errors (PE) beyond ±20% and ±30% was considered superior.

In addition to the prediction-based approaches described above, bootstrap [39] and simulation-based prediction-corrected visual predictive checks (pcVPCs) [40] were conducted to evaluate candidate model predictability. Using Perl modules, 2000 bootstrap datasets were generated by random sampling with replacement [41]. The final popPK model was compared against each of these bootstrap datasets, and all model parameters were estimated.

The dataset was simulated 2000 times for pcVPCs. The 95% confidence intervals (CI) for the median and the 5^th^ and 95^th^ percentiles of the simulations at different times were calculated and compared to the observations, binning automatically.

### 2.6 Nomenclature of targets and ligands

Key protein targets and ligands in this article are hyperlinked to corresponding entries in http://www.guidetopharmacology.org, the common portal for data from the IUPHAR/BPS Guide to PHARMACOLOGY [appropriate reference number], and are permanently archived in the Concise Guide to PHARMACOLOGY 2015/16 [appropriate reference number(s)].

## 3 Results

### 3.1 Patients

Demographic characteristics of the study population are presented in Table 1. A total of 2969 CsA whole-blood measurements were collected from 173 patients after three days post-operation, including 1328 C_0_ and 1641 C_2_. There was a median of 15 CsA observations per patient (range, 2 - 50). No concentrations below the lower quantification limit were included in the analysis.

### 3.2 Population Pharmacokinetic Model Development

#### 3.2.1 Base Model

To compare the potential influence of the structural model on model predictability, a one-compartmental model with first-order absorption and MM empirical formula were used as a structural model to describe CsA PKs. Based on the OFV and the distribution of residuals in the diagnostic plots of the base model, an exponential error model and additive error model were selected to model the random variability for three CMT models and the MM model, respectively.

#### 3.2.2 Covariate Model

The final model parameter estimate results are shown in Table 2. When covariates were included in univariate analysis, neither bodyweight nor other demographic variables provided a significant drop in the OFV (*P* < 0.01).

**Table 2.** Parameter estimates of four popPK models based on different modelling strategies.

| Parameters | *Strategy I:*<br>Linear compartmental model<br>without incorporating DD | *Strategy II:*<br>Linear compartmental model<br>incorporating DD | *Strategy III:*<br>Theory-based nonlinear<br>compartmental model<sup>a</sup> | *Strategy IV:*<br>Nonlinear MM empirical model |  |
| --- | --- | --- | --- | --- | --- |
|  |  |  |  | C<sub>0</sub> | C<sub>2</sub> |
| Objective function value | 32858.4 | 32706.3 | 32617.4 | 11331.9 | 14132.7 |
| K<sub>a</sub> (h<sup>-1</sup>) | 1.25 (fixed) | 1.25 (fixed) | 1.25 (fixed) | / | / |
| CL/F (L h<sup>-1</sup>) | 25.2 (2.0) | 24.6 (1.8) | 14.3 (2.5) | / | / |
| V·(FFM/50)/F (L) | 162.0 (2.8) | 162.0 (2.8) | 254.0 (2.7) | / | / |
| K<sub>m</sub> (ng ml<sup>-1</sup>) | / | / | / | 45.3 (30.9) | 182.0 (20.8) |
| V<sub>m</sub> (mg day<sup>-1</sup>) | / | / | / | 334.0 (1.7) | 329.0 (1.7) |
| Covariate effect on CL/K<sub>m</sub><sup>c</sup> |  |  |  |  |  |
| HCT | -0.404 (12.6) | -0.354 (14.8) | / | / | / |
| POD | 0.00344 (44.8) | 0.0121 (16.4) | 0.0119 (16.1) | 0.336 (38.7) | 0.309 (23.1) |
| DD | / | 0.278 (19.3) | 3.4 (4.2)<sup>b</sup> | / | / |
|  | / | / | 72.8 (26.6)<sup>b</sup> | / | / |
| PD | / | / | 1.08 (1.2) | / | / |
| ω<sub>CL/F</sub> (%) / ω<sub>K<sub>m</sub></sub> (%)<sup>c</sup> | 22.8 (10.4) | 19.9 (11.4) | 19.1 (12.0) | 107.2 (11.8) | 103.4 (10.5) |
| η - shrinkage (%) | 6.3 | 7.7 | 7.9 | 24.0 | 18.4 |
| σ (%/ng ml<sup>-1</sup>)<sup>d</sup> | 34.5 (3.5) | 34.1 (3.2) | 33.8 (3.2) | 32.9 (5.8) | 35.2 (4.7) |
| ε - shrinkage (%) | 3.8 | 3.7 | 3.6 | 9.4 | 8.2 |

Estimates are expressed as estimate (% standard error)
C<sub>0</sub>, pre-dose concentration; C<sub>2</sub>, 2-h post-dose concentration; CL/F, apparent clearance; DD, CsA daily dose; F, the bioavailability relative to 1; FFM, fat-free mass; HCT, haematocrit; K<sub>a</sub>, absorption rate constant; K<sub>m</sub>, steady-state concentrations at half-maximal dose rate; MM, Michaelis-Menten; PD, prednisolone daily dose; POD, postoperative days; V/F, apparent volume of distribution; V<sub>m</sub>, maximum daily dose rate; ω, between subject variability; σ, residual variability
<sup>a</sup> The parameters estimated are expressed as plasma pharmacokinetic parameters in *Strategy III*
<sup>b</sup> 3.4 is the maximum change in CL with increasing CsA daily dose; 72.8mg is the CsA daily dose with half maximum on CL
<sup>c</sup> The parameters estimated in *Strategy I* and *II* are the results on apparent whole blood clearance; the parameters estimated in *Strategy III* are the results on apparent plasma clearance; the parameters estimated in *Strategy IV* are the results on K<sub>m</sub>
<sup>d</sup> Exponential error model is selected for *Strategies I-III* and additive error model is selected for *Strategy IV*

The influence of patients’ body size on CsA disposition was best described by allometric scaling based on fat free mass rather than total body weight for all the PK disposition parameters. After body size, the effect of HCT on CsA clearance was investigated. The OFV substantially declined as HCT was included. *Strategy III*, which accounted for differences in CsA whole-blood concentrations due to haematocrit variation via estimation of C_p_, was superior to *Strategy I and Strategy II* that estimated CsA PK parameters based on whole-blood concentrations alone (△OFV −140.4 vs - 123.6, *P* < 0.001).

Prednisolone dose had no significant influence on CL/*F* when it was investigated empirically in *Strategy I* and *Strategy II*. As prednisolone dosage tapered dramatically in the initial stage of transplantation, the theory-based *Strategy III* tested the effect of prednisolone on *F* based on the hypothesis that the high prednisolone dosage may influence CsA absorption nonlinearly. Therefore, a sigmoid E_max_ model describing an effect of prednisolone dose on *F* (Eq. 4) reduced the OFV by −54.9 (*P* < 0.001) and was superior to the linear model (△OFV −40.8). However, the relative standard error estimated for Pred_50_ was 136.9%, indicating a sparse sampling design that was unable to yield reliable estimates of the nonlinear prednisolone effect on CsA absorption. Thus, the coefficient was introduced to describe this effect. The CL/*F* of patients receiving combined immunosuppressive therapy in the first 15 PODs was 8% higher than the following times (△OFV −40.5, Eq. 9).

A significant increase (119.7%) in CL/*F* was seen as the CsA daily dose (DD) increased from 50 to 600 mg (△OFV-22.9, *P* < 0.001) in *Strategy III*, indicating a nonlinear relationship between DD and CL/*F*. Based on *Strategy I*, DD was also involved in *Strategy II* (△OFV −77.9, *P* < 0.001), indicating a significant model improvement. In addition to these covariate effects, CL/*F* was found to significantly increase with POD in an exponential function for all three CMT models.

For nonlinear *Strategy IV*, as the CsA oral clearance seemed to change with time, the influence of POD on K_m_ was included both for C_0_ (△OFV −611.6, *P* < 0.001) and C_2_ (△OFV −743.3, *P* < 0.001). Then, time factors (A/θ, Eq. 12) were introduced to test whether the MM model with time-variant K_m_ was superior. The weighted residuals (y-axis) showed a clear trend when plotted against the PODs before time factors were added (Fig. S1, *left panel*). This trend was resolved with the time-variant model (Fig. S1, *right panel*).

In final models, all retained covariates caused a significant increase in OFV upon removal. The following were determined to be the final models with CL/*F* or K_m_ covariates.

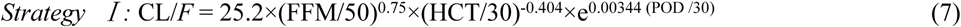

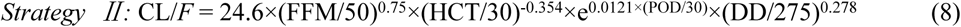

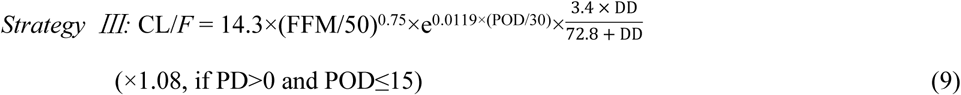

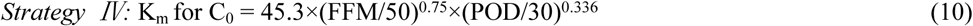

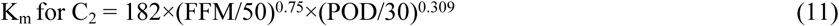

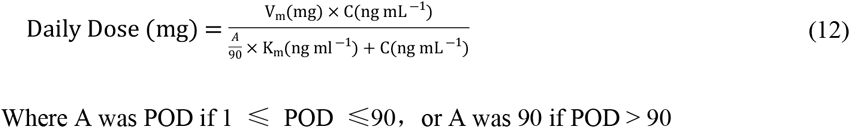

in which the influence scopes were adjusted by their respective median values determined from the dataset.

The estimated PK parameters of the final models were comparable with those of previously published models for CsA in renal transplant recipients. Based on a standard haematocrit value of 30%, the corresponding whole blood apparent clearance of *Strategies I-III* were 25.2 h^-1^, 24.6 h^-1^, and 25.1 l h^-1^, respectively. The MM constant (K_m_) was 45.3 ng/ml for C_0_ and 182 ng/ml for C_2_ one month after transplantation, respectively.

### 3.2.3 Model evaluation

The goodness-of-fit plots for the base and four final models are presented in Fig. S2. Compared to the base model, the final models were greatly improved and showed no structural bias. pcVPCs of the four final models are depicted in Fig. 1. The simulated data corresponded well with the observed data, indicating the lack of significant model misspecifications. Consistently, bootstrap parameter estimates closely matched the mean estimates from the population model, confirming model stability (Table S1).

The F_30_ of the nonlinear MM model showed a remarkable improvement over the other three CMT models, regardless of whether the base (63.4% versus 52.4%) or final (71.1% versus 56.2%, 58.2% and 59.8%) models were used (Fig. 2 and Table 3). The theory-based nonlinear model performed better than the other two CMT models. The predictability improved as nonlinearity was considered.

**Table 3.** A summary of external evaluation.

| Candidate models | MDPE (%) | MAPE (%) | F<sub>20</sub> (%) | F<sub>30</sub> (%) |
| --- | --- | --- | --- | --- |
| Linear compartmental base model | -6.1% | 28.7% | 35.5% | 52.4% |
| Nonlinear MM base model | 9.2% | 20.3% | 48.9% | 63.4% |
| Linear compartmental model without incorporating DD (*Strategy I*) | -3.9% | 26.1% | 40.6% | 56.2% |
| Linear compartmental model incorporating DD (*Strategy II*) | -0.5% | 24.8% | 40.5% | 58.2% |
| Theory-based nonlinear compartmental model (*Strategy III*) | -1.4% | 23.9% | 42.3% | 59.8% |
| Nonlinear MM empirical model (*Strategy IV*) | 6.5% | 16.2% | 59.4% | 71.1% |

DD, Cyclosporine daily dose; F<sub>20</sub> and F<sub>30</sub>, the percentage of prediction error $\leq \pm 20\%$ and $\pm 30\%$ , respectively; MAPE, median absolute prediction error; MDPE, median prediction error; MM, Michaelis-Menten

## 4 Discussion

Empirical investigation of nearly 20 popPK models developed for CsA to aid in the prediction of initial and maintained doses in kidney transplant recipients revealed inconsistent covariate identification. According to our previous investigation [11], the predictability of these empirical CMT models was not precise enough when extrapolated to another clinical centre. Most of these models had not fully considered CsA nonlinear property in modelling processes.

The current study developed four different modelling strategy-based popPK models and compared predictive performances to determine the potential influence of nonlinearity and the functional forms of covariates on model predictability. The conclusion that predictability improved as nonlinearity was considered in the CsA modelling process was demonstrated in this large cohort study. Incorporating covariates based on a fundamental understanding of PK processes can help to improve model predictability. The predictive performance of the nonlinear MM empirical model was much better than that of the compartmental models, indicating that other sources of CsA PK nonlinearity should be considered in further analyses.

Unlike empirical covariate selection, theory-based covariate selection allows the incorporation of relationships linking parameters and covariates based on a fundamental understanding of PK processes rather than on the available data alone [22]. In contrast to the empirical approaches in previous studies, covariates in *Strategy III* were investigated based on theoretical mechanisms, and the potential mechanisms for CsA PK nonlinearity were determined by pop PK analysis. Allometric scaling to fat free mass, saturated distribution into red blood cells, and the influence of CsA daily dose on metabolism were all incorporated into the final model based on the fundamental understanding of PK processes. The change in bioavailability caused by tapering the prednisolone dose in the initial stage of transplantation, other pathophysiological factors, and drug-drug interactions were also investigated. Most of these factors contributed to CsA PK nonlinearity.

The influence of haematocrit on the CsA PK process was primarily considered because approximately 58% of circulating CsA was bound to red blood cells [42]. According to the work of Lemaire *et al*. [34] and Legg *et al*. [32], CsA binding to plasma components is linear, while binding to erythrocytes is nonlinear. Haematocrit is reduced in transplant patients and occurs at a lower level in the early postoperative period [42]. To reduce the confounding effect of haematocrit variability in predicting CsA concentrations, we hypothesised that the plasma concentration is proportional to the unbound CsA concentration, and the PK parameters of *Strategy III* are established from implicit plasma concentrations rather than whole blood concentrations. Based on a standard haematocrit value of 30%, the corresponding whole-blood apparent clearance (CL_wb_/*F*) was 25.1 l h^-1^ This is comparable to previously published values (17.0-50.4 l h^-1^) [11] and consistent with the results of the linear compartmental model [43].

To date, almost all published CsA popPK models determined the haematocrit effect on CsA PK processes through an empirical approach, such as *Strategies I* and *II*, and only two studies retained this covariate in the final model [44, 45]. This phenomenon was inconsistent with the CsA distribution mechanism, which may influence model predictability. Furthermore, CsA TDM is based on whole-blood concentrations, which have a very variable relationship with unbound concentrations. The unbound concentration is considered more closely related to accurately determining efficacy or side effects [46]. Therefore, this should be the focus of future studies.

The obvious improvement in predictability observed when the daily dose was incorporated (*Strategy I* versus *Strategy II*) illustrated apparent clearance dose dependency. *In vitro*, CsA is a moderately strong CYP3A4- and strong P-gp inhibitor [14, 15, 47]. Based on formation of the M1 metabolite as a CYP3A4 probe, hepatic CYP3A4 activity decreases by up to 26% in the presence of CsA [48]. The inhibitory effects of CsA on CYP3A4 and P-gp can lead to autoinhibition and dose-dependent PKs, which may also contribute to CsA nonlinearity [47, 49]. Therefore, in *Strategy III*, the CsA daily dose was included nonlinearly; CL/*F* increased from 12.7 l h^-1^ to 27.9 l h^-1^ as the CsA daily dose changed from 50 mg day^-1^ to 600 mg day^-1^, demonstrating that increasing the CsA dose produces a disproportional increase in CsA exposure.

In healthy volunteers administered a single oral dose, the CsA PK is normally considered linear [8]. However, physiology disorders, co-administrated drugs, and other intrinsic factors in clinical testing may influence CsA PK behaviour. In the present analysis, the bioavailability change caused by tapering the prednisolone dose in the early PODs was considered by initially incorporating the effect of prednisolone dose on *F* nonlinearly. However, the sparse sampling design could not yield reliable estimates of the nonlinear prednisolone effect on CsA absorption. An 8% clearance increase was observed when a higher steroid dosage was received in the initial stage of transplantation; this phenomenon should be further elucidated. Variability from concomitant calcium channel blockers was considered but no significant effect was found.

According to previous findings, WT and POD are the most recognised covariates in empirical building studies [11]. WT-relevant factors were considered in the present study and fat free mass was included. It is reasonable to accept fat-free mass as a predictor of CsA clearance because fat mass is not expected to directly influence metabolic capacity [25]. POD, as a surrogate for many time-dependent factors, was included in the final model. Some factors that are known to systematically change with POD, such as HCT and DD, were also investigated. Other currently unknown POD-related factors should be explored in future studies.

The retrospective observational study design did not allow confirmation of whether patients adhered to their prescribed dosage regimen, and no intensive sampling was available in this study. The sparse sampling design could not yield reliable estimates of the nonlinear prednisolone effect on CsA absorption.

By comparing the predictive performance of four popPK models based on different modelling strategies, we found that incorporating nonlinear properties is likely to be a promising approach for improving CsA model predictability. Saturated distribution into red blood cells, and auto-inhibition that arose from the inhibitory effects of CsA on *CYP3A4*/P-gp and CsA-prednisolone drug interaction may be the main sources of CsA PK nonlinearity.

## Data Availability

The datasets used and/or analyzed during the current study are available from the corresponding author on reasonable request.

## 5 Compliance with Ethical Standards

### Funding

This work was supported in part by grants from the National Natural Science Foundation of China (No. 81 573 505 and 81 072 702), the “2016 Key Clinical Program of Clinical Pharmacy”, and “Weak Discipline Construction Project” (No. 2016ZB0301-01) of Shanghai Municipal Health and Family Planning Commission.

### Ethical Approval

All procedures involving human participants were in accordance with the ethical standards of the institutional and/or national research committee and with the 1964 Helsinki Declaration and its later amendments or comparable ethical standards.

### 6 Contributors

JJM and ZJ participated in the research design. XYQ, MKZ and MZ helped acquire the evaluated data. JJM performed the research and analysed the data. JJM and ZJ drafted the manuscript, which was revised and approved by all the authors. There are no other relationships or activities that could appear to have influenced the submitted work.

#### 7 Acknowledgments

The authors are grateful to Dr. Chen-Yan Zhao of the University of Uppsala for her invaluable advice, and Han-Chao Chen of Huashan Hospital for his support during data editing.

### 8 Data availability statement

The data that support the findings of this study are available from the corresponding author upon reasonable request.

---

**Fig. 1.**
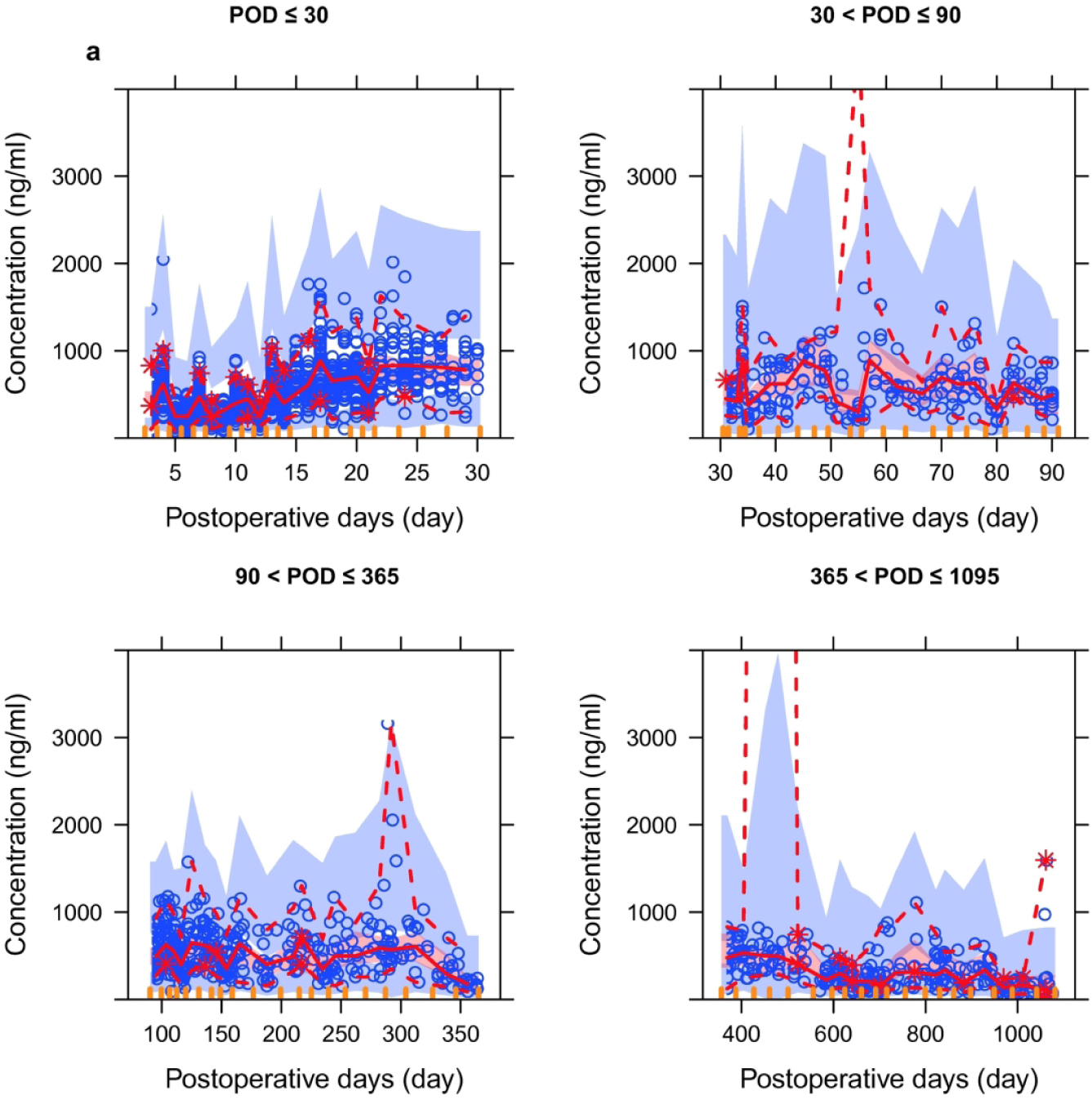
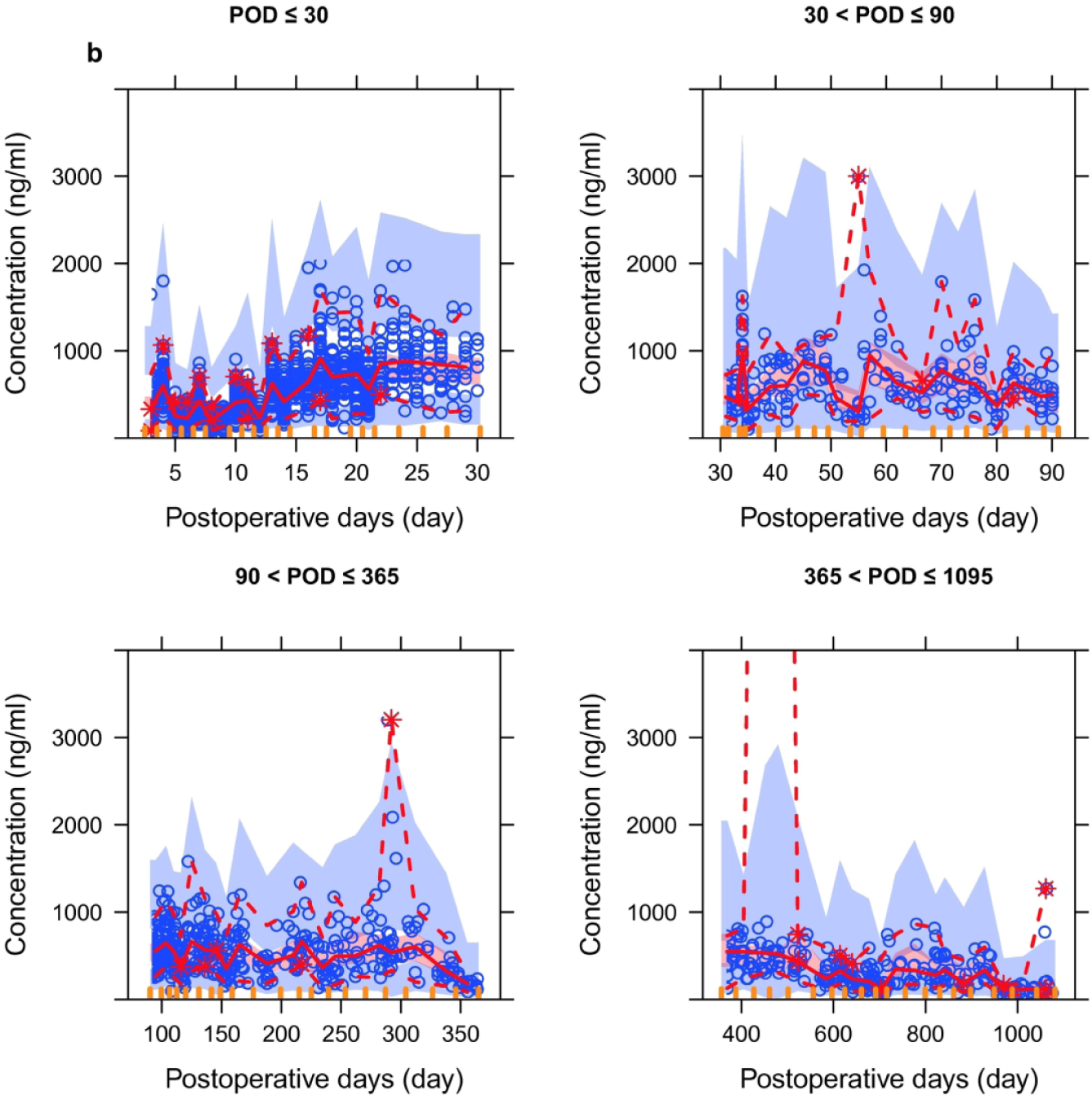
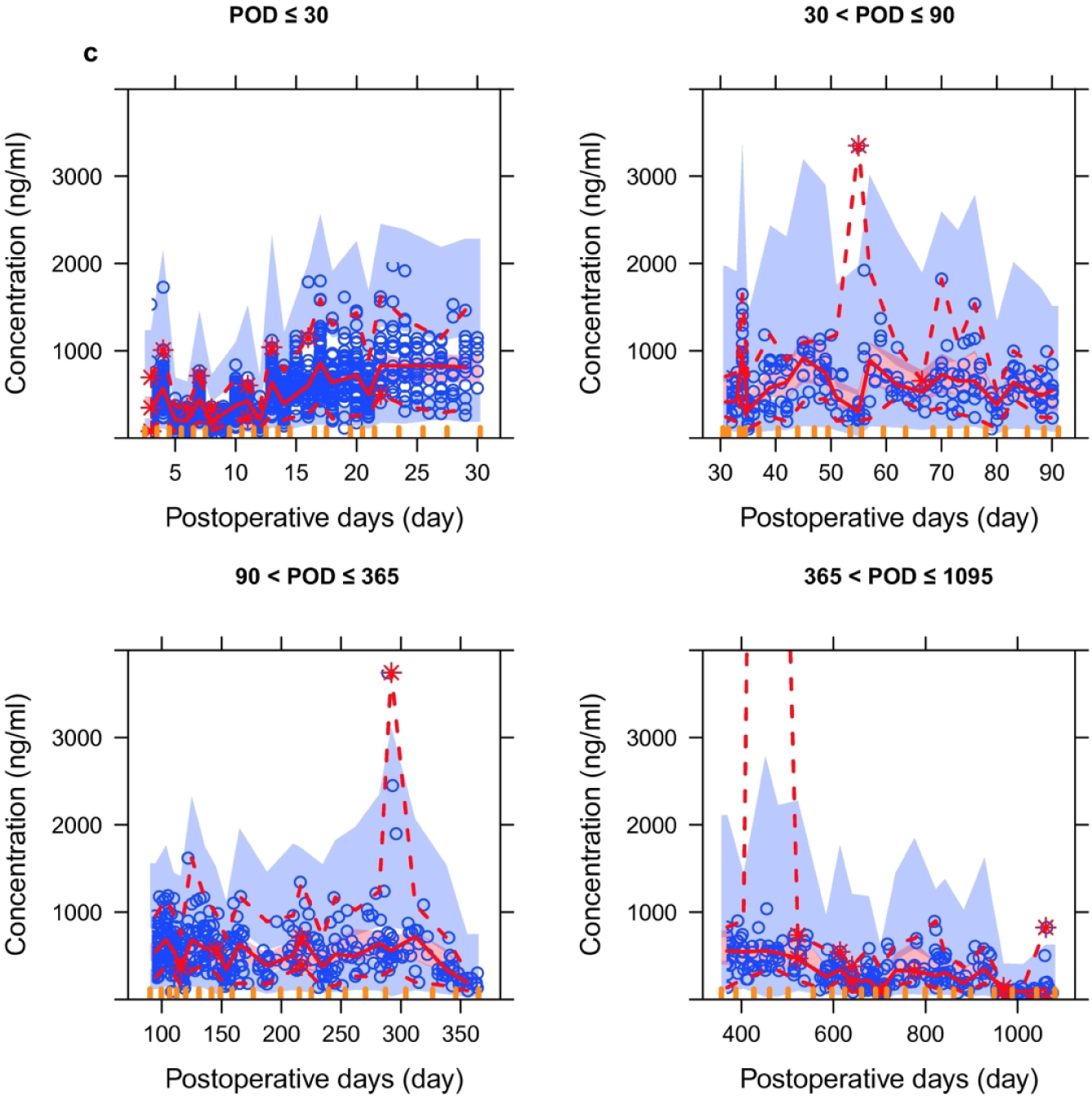
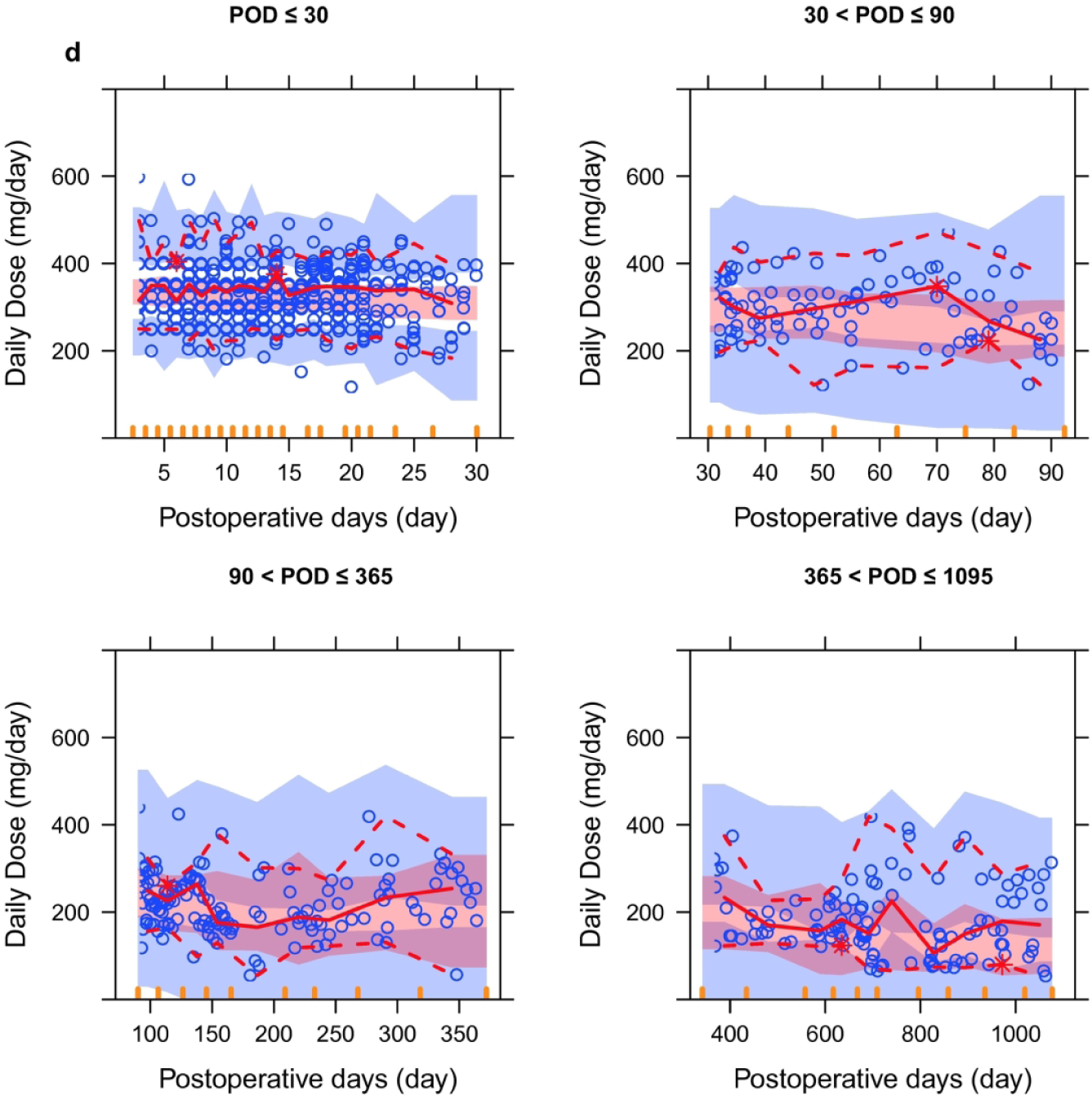
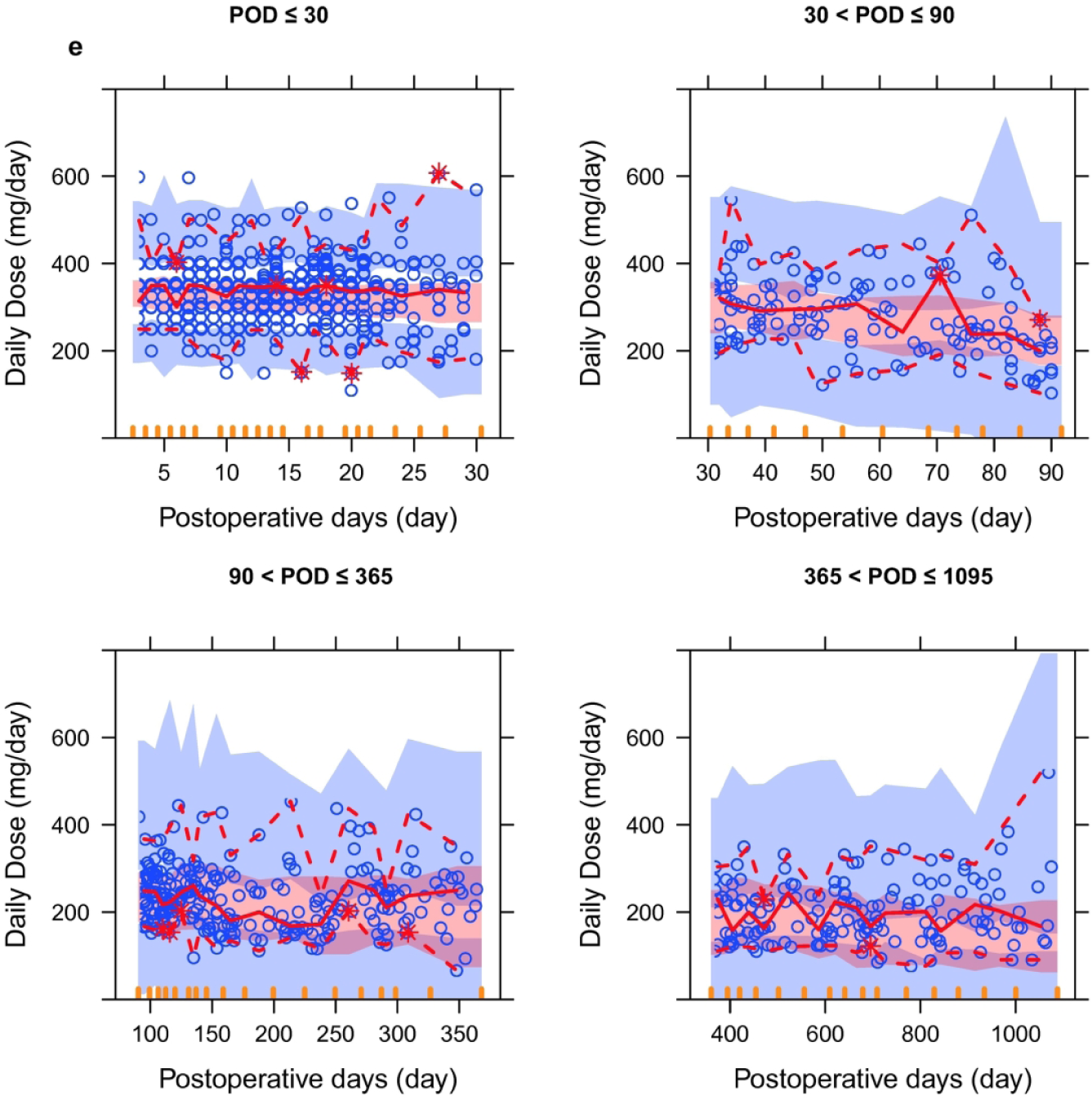
Prediction-corrected visual predictive checks (pcVPCs) stratified on postoperative days (PODs) for four established final models, based on 2000 simulations. The *red solid line* connects median observed values per bin, the *red dashed lines* connect the 5^th^ and 95^th^ percentiles of the observations. The *blue areas* are the 95 % confidence interval of the 5^th^ and 95^th^ percentiles. The *red area* indicates the confidence interval of the median. **a** pcVPC of linear compartmental model without incorporating daily dose, **b** pcVPC of linear compartmental model incorporating daily dose, **c** pcVPC of theory-based nonlinear compartmental model, **d** pcVPC of nonlinear Michaelis-Menten empirical model for pre-dose concentrations, **e** pcVPC of nonlinear Michaelis-Menten empirical model for 2-h post-dose concentrations. 177×177mm (300 × 300 DPI)

**Fig. 2.**
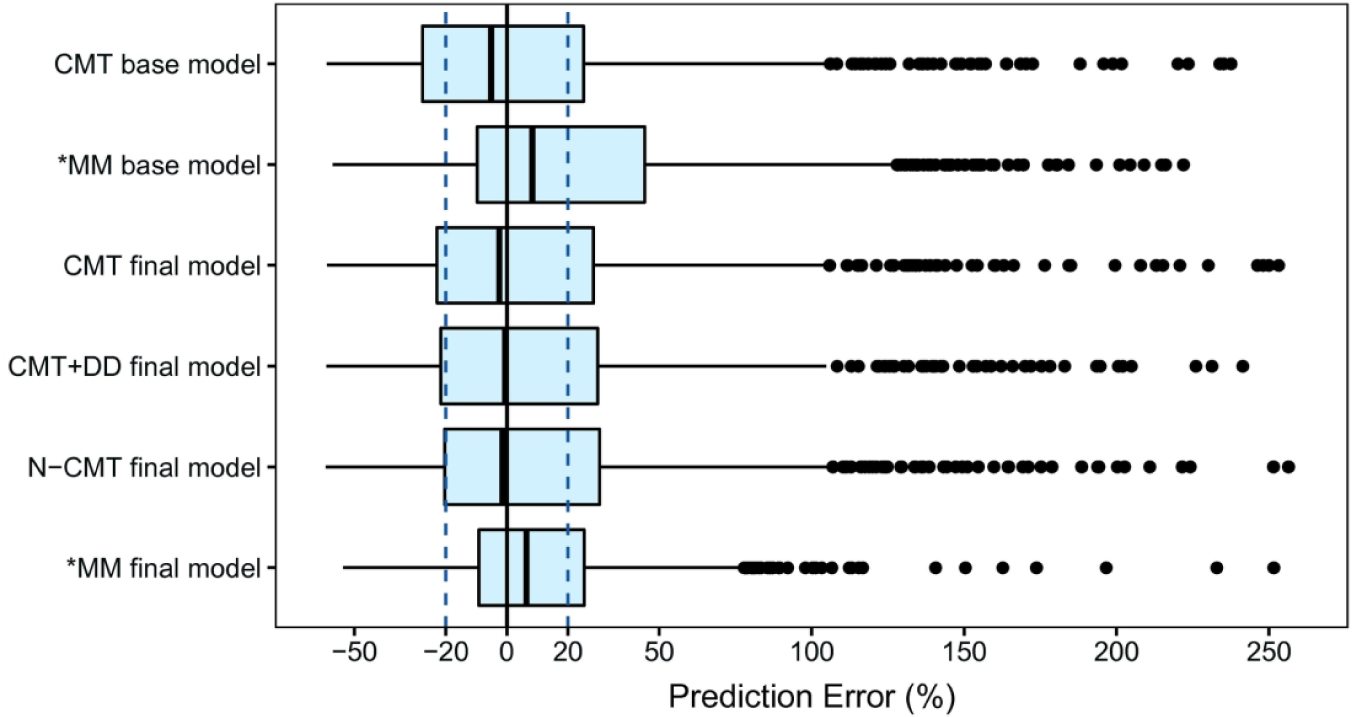
Box plots of the prediction error (PE%) for the four established final models. *Black solid lines* and *blue dotted lines* are reference lines indicating PE% of 0% and ± 20%, respectively. *CMT* linear compartmental model, *DD* cyclosporine daily dose, *MM* Michaelis-Menten model, *N-CMT* nonlinear compartmental model. Models with an asterisk (*) were developed based on the Michaelis-Menten model. 181×94mm (300 × 300 DPI)

## Supporting information

Additional Supporting Information may be found in the online version of this article at the publisher’s web-site:

***Supplementary Text S1*** Target concentrations of CsA therapeutic monitoring

***Supplementary Text S2*** The semi-mechanistic model used to predict fat-free mass

***Supplementary Text S3*** The derivation process of formula used to predict CsA plasma concentrations

***Table S1*** A summary of parameters estimated by four final models using 2,000 bootstrap runs

**Figure S1.**
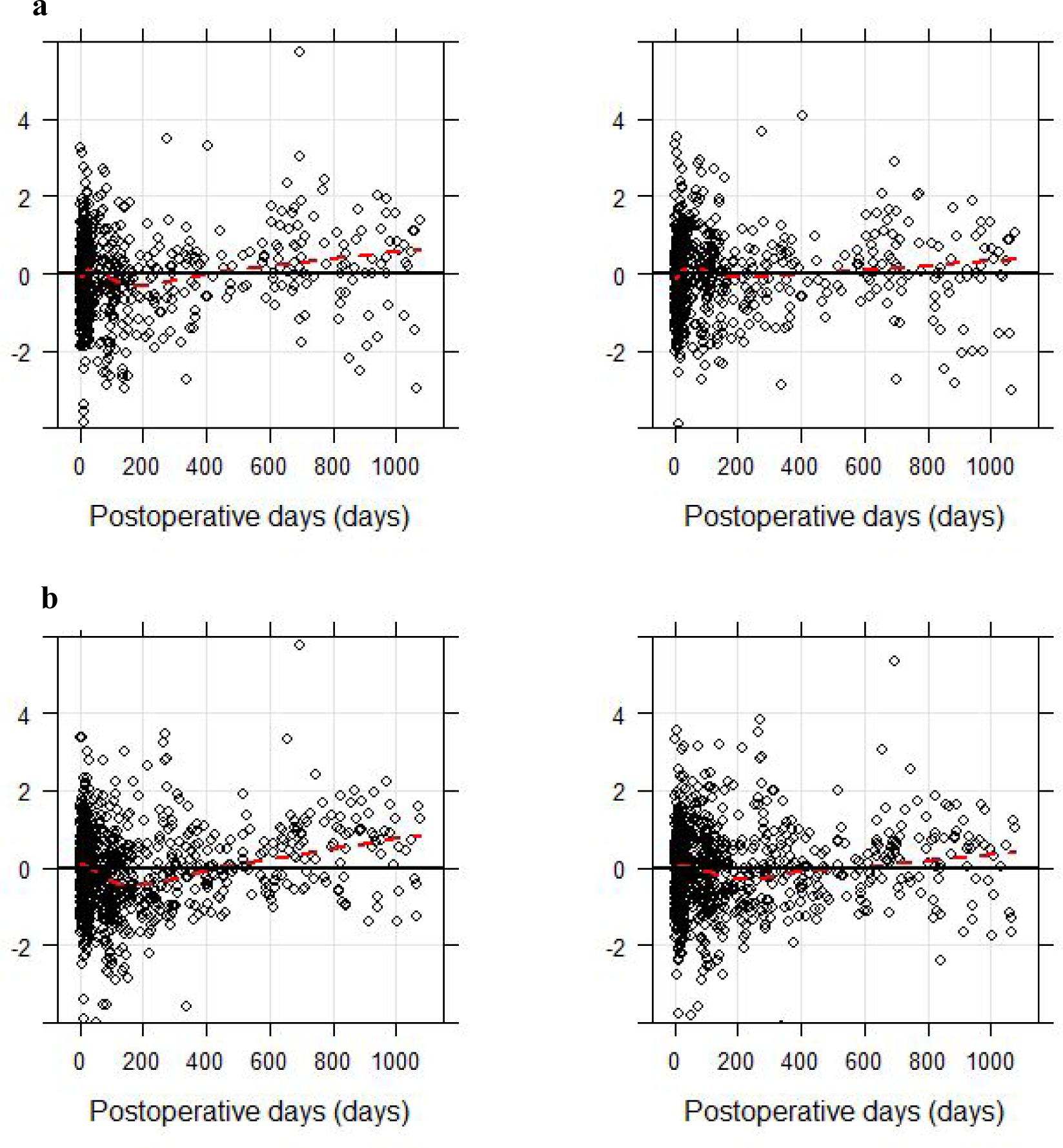
Residual plot of the time-invariant (*left panel*) and the time-variant (*right panel*) Michaelis-Menten model. The conditional weighted residuals (CWRES) are plotted against the postoperative days. **a** the results of Michaelis-Menten model for pre-dose concentrations, **b** the results of Michaelis-Menten model for 2-h post-dose concentrations. *Red lines* represents the LOESS smoothing.

**Figure S2.**
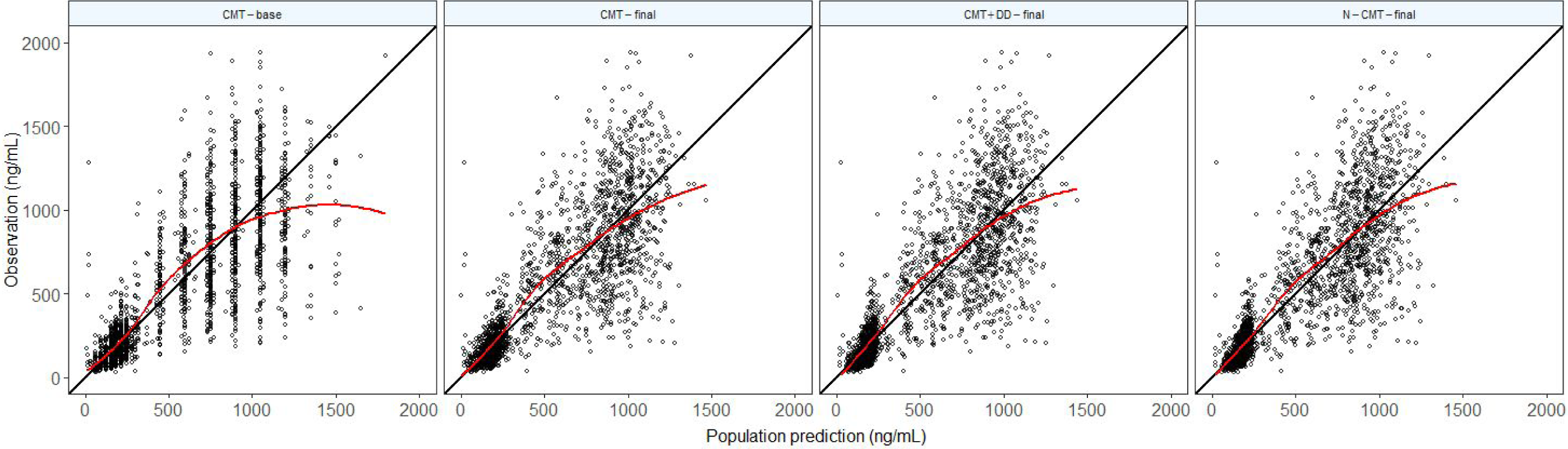
**a-h** Goodness-of-fit plots for base model and four final models. **a**,**e** population prediction versus observation, **b**,**f** individual prediction versus observation, **c**,**g** population prediction versus conditional weighted residuals (CWRES), **d**,**h** postoperative days versus CWRES. **a-d** the results of compartmental models, **e-h** the results of Michaelis-Menten models. *Solid lines* in **a**,**b**,**e**,**f** represent lines of identity, *Red lines* represent LOESS smoothing. *CMT* compartmental model, *CMT+DD* linear compartmental model incorporating daily dose, *N-CMT* theory-based nonlinear compartmental model, *MM-C*_*0*_ nonlinear Michaelis-Menten empirical model for pre-dose concentrations, *MM-C*_*2*_ nonlinear Michaelis-Menten empirical model for 2-h post-dose concentrations.

## Electronic Supplementary Material

### Supplementary Text S1 Target concentrations of CsA therapeutic monitoring

Target C_0_ values were 200 - 350 ng ml^-1^ in the first month, 150 - 300 ng mL^-1^ during months 1 to 3, 100 - 250 ng mL^-1^ during months 3 to 12, and 50 - 100 ng mL^-1^ thereafter; target C_2_ values were 1000 - 1500 ng mL^-1^ in the first month, 800 - 1200 ng mL^-1^ during months 1 to 3, 600 - 1000 ng mL^-1^ during months 3 to 12, and 400 - 600 ng mL^-1^ thereafter.

### Supplementary Text S2 The semi-mechanistic model used to predict fat-free mass (FFM)

The semi-mechanistic model be used to predict FFM as follows [1]:

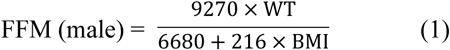

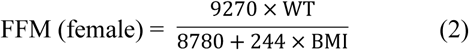

Where WT is measured in kilograms, BMI is calculated following the formula: BMI = weight (kg)/height^2^ (m^2^), *BMI* body mass index, *WT* body weight

#### Supplementary Text S3 The derivation process of formula used to predict CsA plasma concentrations (C_u_)

Assuming that C_u_ is the same both inside and outside the erythrocyte, then the concentration of drug in packed erythrocytes (C_e_) is the combine of C_u_ and the concentration of drug bound to material in packed erythrocytes (C_be_).

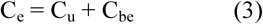

It is assumed that an equilibrium exists between C_u_ and drug bound to a single-site binding material associated with the erythrocyte,

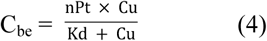

Where P_t_ is the total concentration of binding material in packed erythrocytes, K_d_ is the equilibrium dissociation constant, and n is the number of binding sites per unit of binding material.

Combining Eqs. (3) and (4) gives

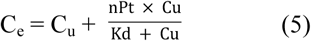

If C_wb_ is the total-blood concentration of drug, then by mass balance and considering a unit volume,

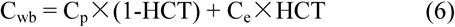

If the function unbound (fu) is independent of C_p_, so that C_u_ = fu × C_p_. With the parameter values estimated with real-world data before (fu = 0.0727, nP_t_ = 2562 μg/L, K_d_ = 67 μg/L) [2], C_wb_ was related to C_p_ and haematocrit.

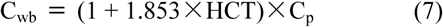

**Supplementary Table S1.** A summary of parameters estimated by four final models using 2,000 bootstrap runs

| Parameters | *Strategy I:*<br>Linear compartmental model<br>without incorporating DD |  | *Strategy II:*<br>Linear compartmental model<br>incorporating DD |  | *Strategy III:*<br>Theory-based nonlinear<br>compartmental model<sup>a</sup> |  | *Strategy IV:*<br>Nonlinear MM empirical model |  |  |  |
| --- | --- | --- | --- | --- | --- | --- | --- | --- | --- | --- |
|  | Mean | 2.5<sup>th</sup>-97.5<sup>th</sup><br>percentiles | Mean | 2.5<sup>th</sup>-97.5<sup>th</sup><br>percentiles | Mean | 2.5<sup>th</sup>-97.5<sup>th</sup><br>percentiles | C<sub>0</sub><br>Mean | 2.5<sup>th</sup>-97.5<sup>th</sup><br>percentiles | C<sub>2</sub><br>Mean | 2.5<sup>th</sup>-97.5<sup>th</sup><br>percentiles |
| K<sub>a</sub> (h<sup>-1</sup>) | 1.25 (fixed) | / | 1.25 (fixed) | / | 1.25 (fixed) | / | / | / | / | / |
| CL/F (L h<sup>-1</sup>) | 25.2 | 24.4-26.4 | 24.6 | 23.8-25.6 | 14.5 | 11.4 | / | / | / | / |
| V·(FFM/50)/F (L) | 162.0 | 149.2-167.5 | 162.0 | 150.1-167.9 | 254 | 17.2 | / | / | / | / |
| K<sub>m</sub> (ng ml<sup>-1</sup>) | / | / | / | / | / | / | 45.8 | 20.1-70.4 | 183.8 | 107.6-257.1 |
| V<sub>m</sub> (mg day<sup>-1</sup>) | / | / | / | / | / | / | 334.1 | 322.3-345.3 | 328.8 | 318.2-338.9 |
| Covariate effect on CL/K<sub>m</sub><sup>c</sup> |  |  |  |  |  |  |  |  |  |  |
| HCT | -0.406 | -0.513--0.306 | -0.355 | -0.477--0.266 | / | / | / | / | / | / |
| POD | 0.00343 | -0.0003-0.006 | 0.0122 | 0.006-0.014 | 0.0119 | 0.008-0.016 | 0.347 | 0.079-0.592 | 0.315 | 0.166-0.452 |
| DD | / | / | 0.282 | 0.116-0.335 | 3.4<sup>b</sup> | 2.8-4.0 | / | / | / | / |
|  | / | / | / | / | 74.6<sup>b</sup> | 33.0-112.7 | / | / | / | / |
| PD | / | / | / | / | 1.08 | 1.05-1.10 | / | / | / | / |
| ω<sub>CL/F</sub> (%) / ω<sub>K<sub>m</sub></sub> (%)<sup>c</sup> | 22.9 | 15.1-25.7 | 20.0 | 14.0-23.6 | 19.0 | 13.9-23.2 | 107.8 | 75.5-131.3 | 103.6 | 78.0-123.9 |
| η - shrinkage (%) | / | / | / | / | / | / | / | / | / | / |
| σ (%/ng ml<sup>-1</sup>)<sup>d</sup> | 34.6 | 30.9-35.9 | 34.1 | 30.8-35.3 | 33.7 | 31.6-35.7 | 32.9 | 29.1-36.4 | 35.2 | 31.9-38.3 |
| ε - shrinkage (%) | / | / | / | / | / | / | / | / | / | / |

C<sub>0</sub>, pre-dose concentration; C<sub>2</sub>, 2-h post-dose concentration; CL/F, apparent clearance; DD, CsA daily dose; F, the bioavailability relative to 1; FFM, fat-free mass; HCT, haematocrit; K<sub>a</sub>, absorption rate constant; K<sub>m</sub>, steady-state concentrations at half-maximal dose rate; MM, Michaelis-Menten; PD, prednisolone daily dose; POD, postoperative days; V/F, apparent volume of distribution; V<sub>m</sub>, maximum daily dose rate; ω, between subject variability; σ, residual variability.
<sup>a</sup> The parameters estimated are expressed as plasma pharmacokinetic parameters in *Strategy III*
<sup>b</sup> 3.4 is the maximum change in CL with increasing CsA daily dose; 74.6mg is the CsA daily dose with half maximum on CL
<sup>c</sup> The parameters estimated in *Strategy I* and *II* are the results on apparent whole blood clearance; the parameters estimated in *Strategy III* are the results on apparent plasma clearance; the parameters estimated in *Strategy IV* are the results on $K_m$
<sup>d</sup> Exponential error model is selected for *Strategies I-III* and additive error model is selected for *Strategy IV*

## Electronic Supplementary Material

**Supplementary Fig. S2.**
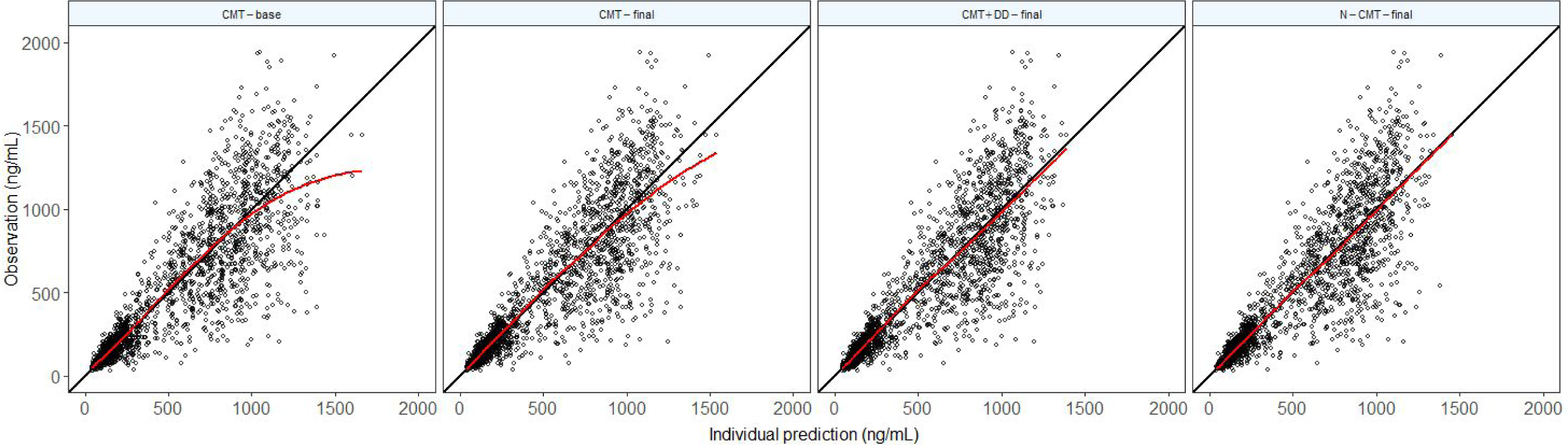

**Supplementary Fig. S2.**
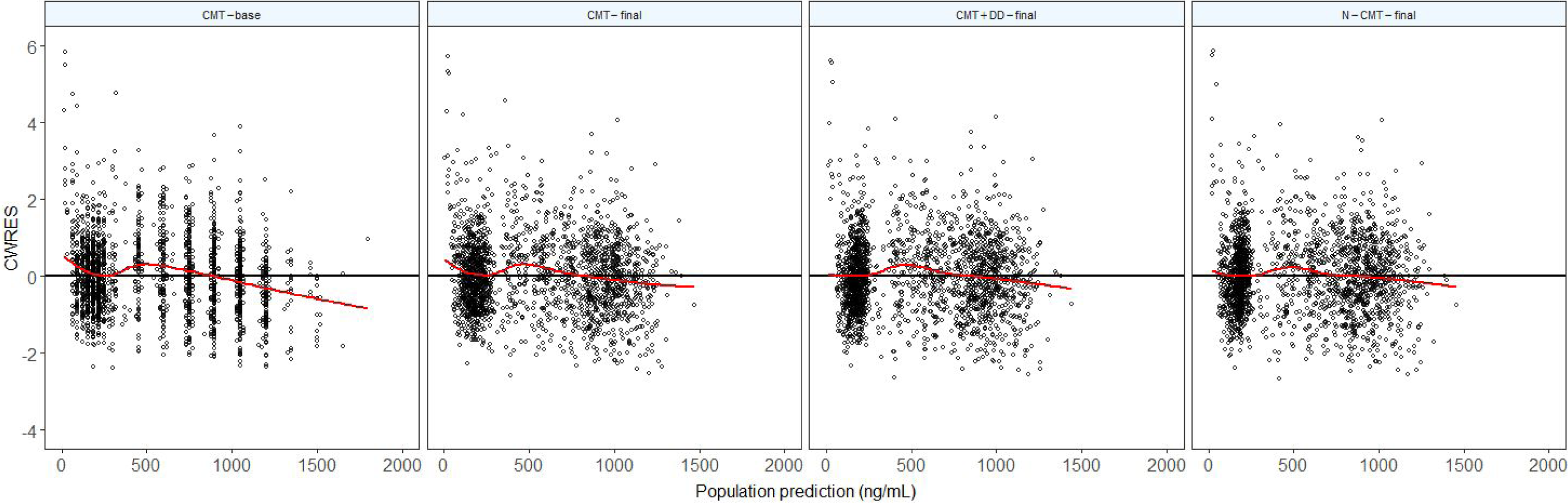

**Supplementary Fig. S2.**
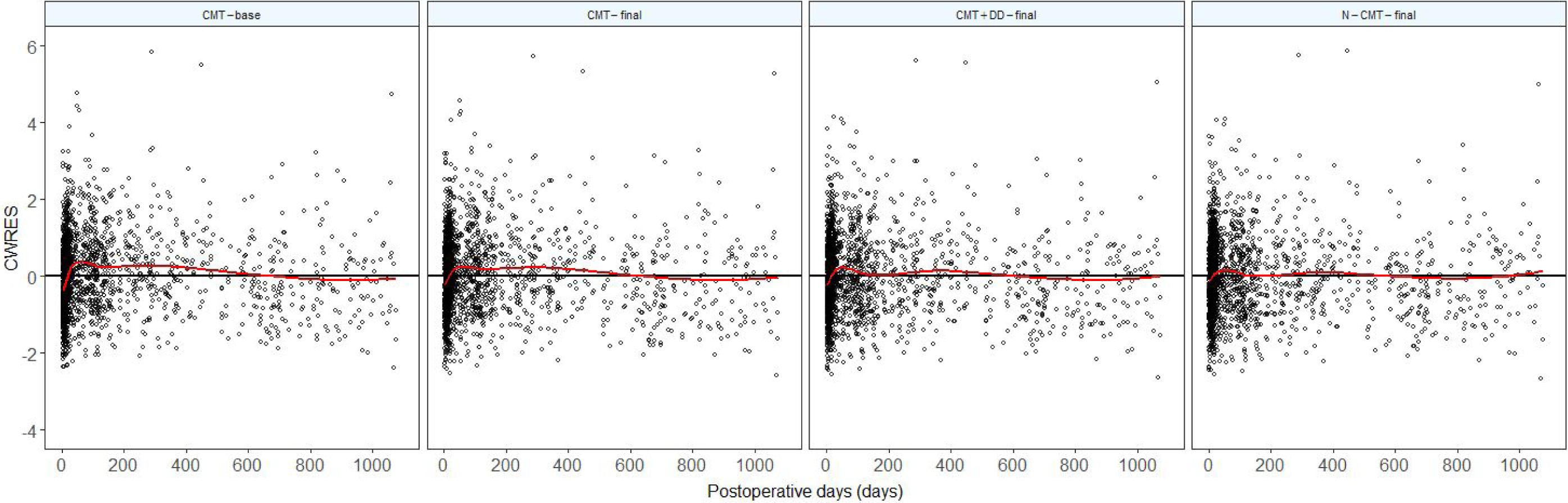

**Supplementary Fig. S2.**
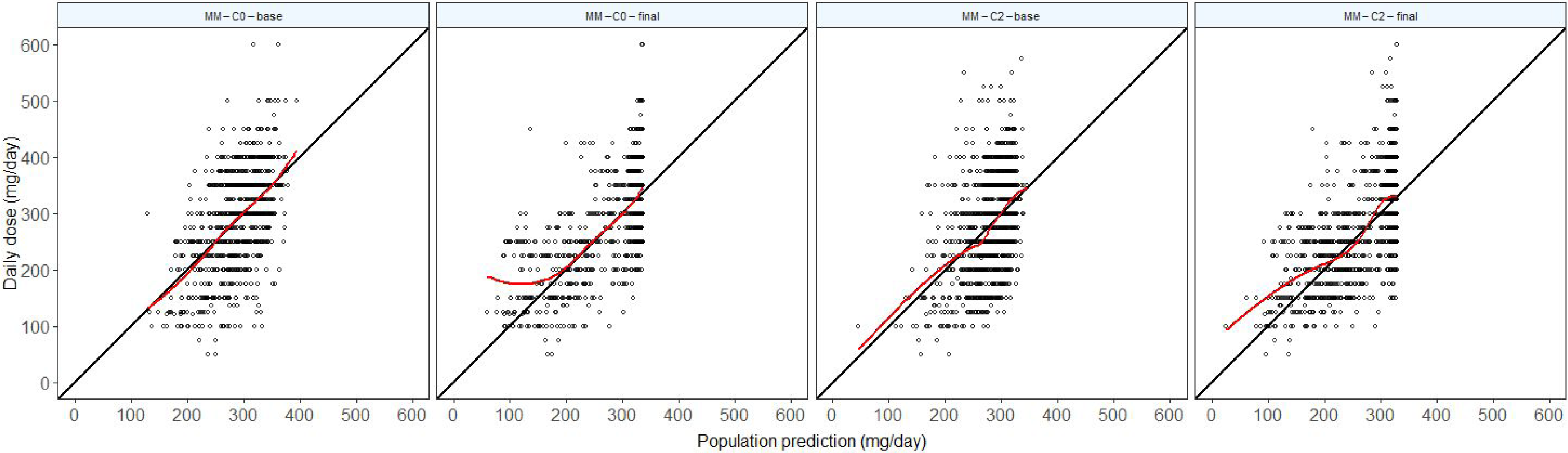

**Supplementary Fig. S2.**
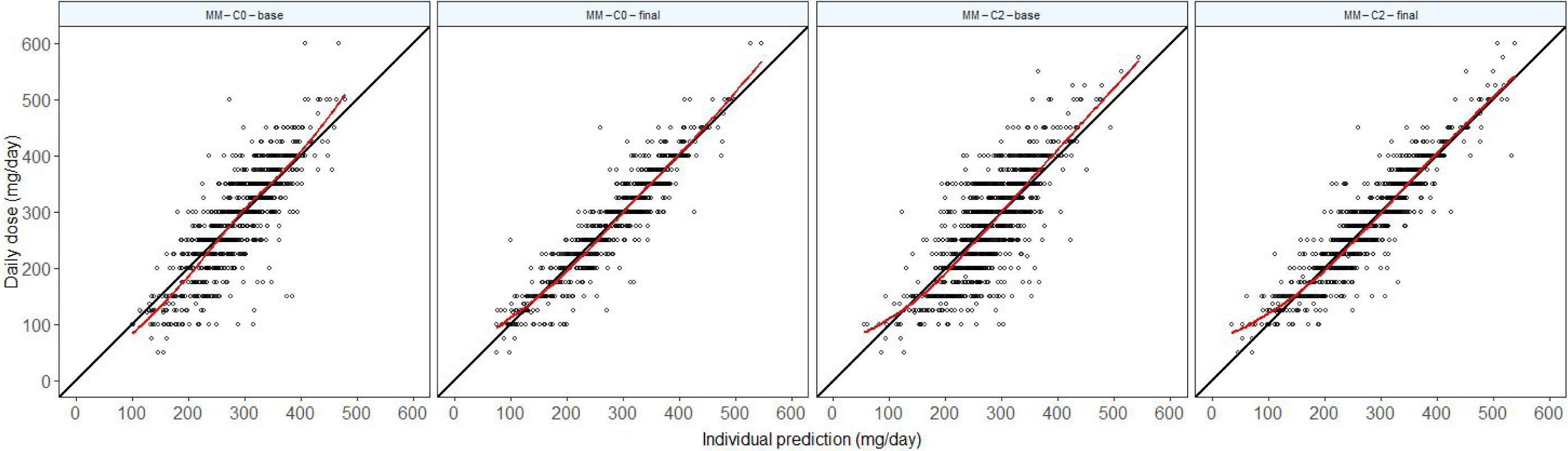

**Supplementary Fig. S2.**
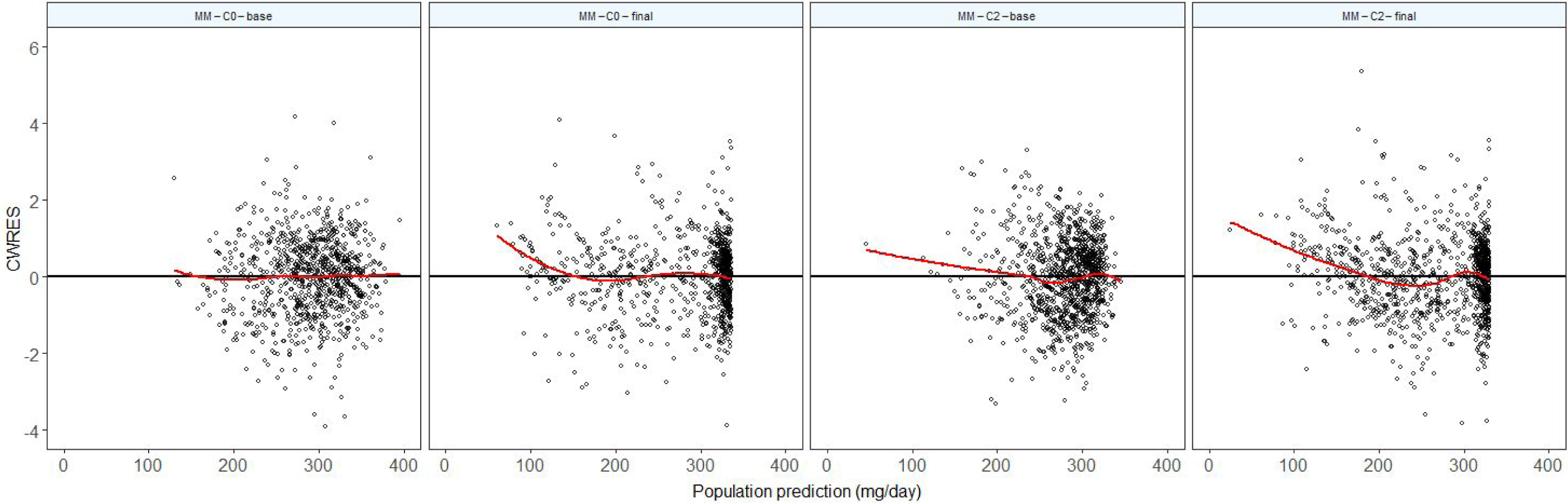

**Supplementary Fig. S2.**
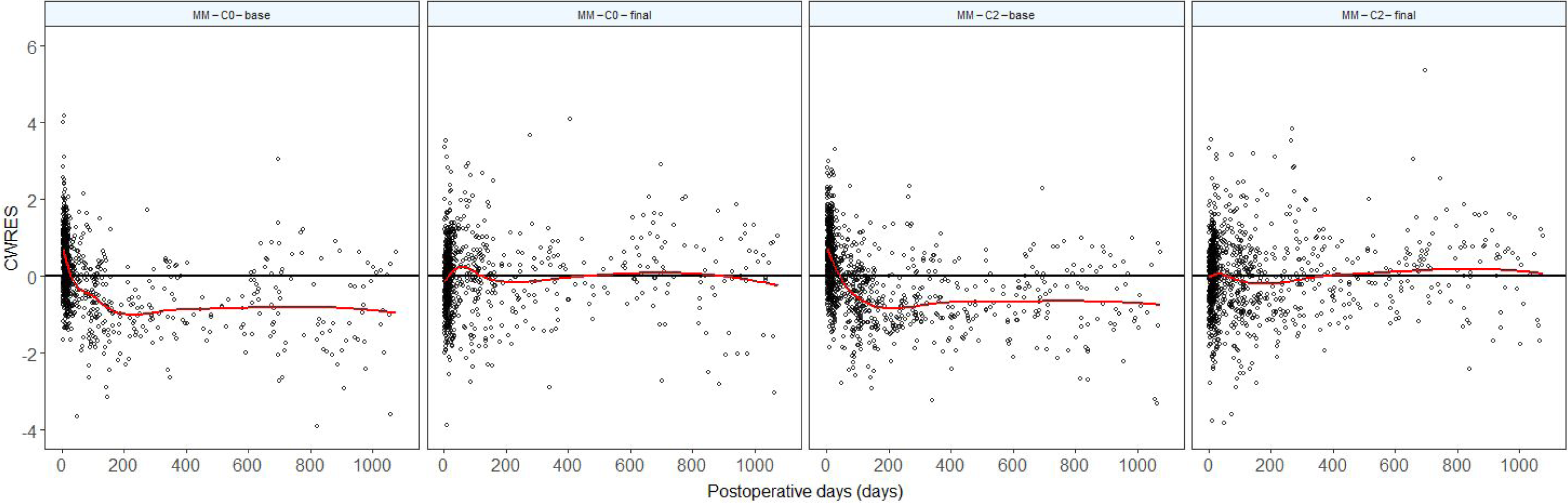

